# A digital twin model incorporating generalized metabolic fluxes to predict chronic kidney disease in type 2 diabetes mellitus

**DOI:** 10.1101/2023.09.22.23295944

**Authors:** Naveenah Udaya Surian, Arsen Batagov, Andrew Wu, Wen Bin Lai, Yan Sun, Yong Mong Bee, Rinkoo Dalan

**Author notes:** Contributing authors.

## Abstract

One of the biggest complication in diabetes patients is chronic kidney disease (CKD), making it a tremendous burden on the country’s public healthcare system. We have developed HealthVector Diabetes (HVD), a digital twin model that leverages generalized metabolic fluxes (GMF) to proficiently predict the onset of CKD and facilitate its early detection. Our HVD GMF model utilized commonly available clinical and physiological biomarkers as inputs for identification and prediction of CKD. We employed four diverse multi-ethnic cohorts (n=7072): one Singaporean cohort (EVAS, n=289) and one North American cohort (NHANES, n=1044) for baseline CKD identification, and two multi-center Singaporean cohorts (CDMD, n=2119 and SDR, n=3627) for 3-year CKD prediction. We developed one identification model and two prediction models (with complete or incomplete parameters). The identification model demonstrated strong performance with an AUC ranging from 0.80 to 0.82. For prediction, with incomplete parameters, we achieved an AUC of 0.75, while the complete parameter model achieved an improved AUC of 0.86. Our model also effectively stratifies patients into low, moderate, and high-risk categories, with the high-risk category having the highest proportion (53.3-62.9%) of patients with CKD. Our method also reveals metabolic health profile differences among patient subgroups at baseline, indicating that patient subgroups who develop future CKD exhibit more deteriorated profiles compared to future non-CKD patients. Furthermore, we also show that GMF-based clustering reveals distinct metabolic profile differences that act as drivers for CKD progression, and the distance between different patient clusters can be used to map patient health trajectories. Our HVD GMF digital twin model has the ability to identify patients with baseline CKD and predict future CKD within a 3-year time frame. Furthermore, our approach enables risk stratification, sub-grouping and clustering based on metabolic health profiles, positioning our model as a valuable clinical application tool for healthcare practitioners.

## 1 Introduction

Diabetes has emerged as a prominent global health crisis in the 21st century, with the lifespan of patients estimated to be reduced by 12 years due to vascular complications [1]. In a multiethnic population as Singapore, the prevalence of diabetes has experienced a twofold increase over the past four decades, and it also has the highest global prevalence of diabetic kidney failure [2]. Among non-fatal complications of diabetes, end-stage renal disease requiring dialysis stands out as a significant driver of healthcare costs [3]. It is therefore crucial to detect chronic kidney disease (CKD) early in its course, before reaching a non-reversible state.

A digital twin can be defined as a direct digital representation of an individual based on the individuals own comprehensive biological data [4]. Using this concept, a virtual replica of the vascular system in the individual can be represented based on a mechanistic model [5]. These digital twin models can then be used to simulate progression of chronic diseases and to predict probable trajectories of disease development [6]. Moreover, these models can inform clinicians how altering specific mechanistic components could change disease trajectories, thereby leading patients towards less severe disease states.

In recent years, digital twin technology has gained popularity in the healthcare sector, where, a PubMed trend search on the keyword “digital twin” revealed a ten-fold increase in the last 5 years. Different techniques, measurements and applications can be used to build a patient’s digital twin. In this paper, we utilize generalized metabolic fluxes (GMFs) to construct personalized digital twins of patients and study the trajectory of microvascular complications leading to chronic kidney disease (CKD) [6]. We termed this proprietary technology as HealthVector Diabetes (HVD). The HVD GMF analysis allows us to represent the long-term changes in the rates of metabolic processes in the body as a mathematical model of personalized metabolic rates. The parameters of the model are obtained as a best fit to the observed clinical and physiological measurements of an individual patient. Our GMF-based digital twin model is used here to identify the disease states of patients at baseline and predict the occurrence of CKD within a three-year period.

## 2 Results

### 2.1 Data set characteristics

The table below provides a detailed summary of the main characteristics of each data set (Table 1). Although the four data sets shared many similarities, several differences were observed. Notably, the EVAS dataset included younger patients (54 [11.1]) with higher levels of glycated hemoglobin (HbA1c) (8.6 [1.8]) compared to those in the CDMD dataset (57 [12.4], 8.0 [1.8]), NHANES dataset (59 [11.9], 7.6 [1.9]), and SDR dataset (61 [11.0], 7.4 [1.6]). The NHANES data set had slightly lower systolic blood pressure (SBP) (130.3 [18.6]), higher serum creatinine (85.0 [56.3]), and body mass index (BMI) (32.5 [7.5]) values compared to the EVAS dataset (133.2 [14.8], 27.7 [5.0], 74.2 [26.9]), the CDMD data set (133.8 [17.9], 26.8 [5.6], 74.0 [22.7]) and the SDR data set (132.1 [15.1], 26.6 [5.5], 70.8 [23.3]).

**Table 1.** Study population characteristics of the EVAS, NHANES, CDMD and SDR data sets.

| Analysis | Identification of CKD |  | Prediction of CKD |  |
| --- | --- | --- | --- | --- |
| Type | Identification model | Identification model | Incomplete inputs | Complete inputs |
| Cohort | EVAS | NHANES | CDMD | SDR |
| Number of patients (Number of CKD positive patients at baseline/future) | 289 (96) | 1044 (360) | 2112 (719) | 3627 (1413) |
| Gender (M(%):F(%)) | 144 (49.8):145 (50.2) | 534 (51.1):510 (48.9) | 1116 (52.8):996 (47.2) | 1917 (52.9):1710 (47.1) |
| Age (Mean, SD) | 54 (11.1) | 59 (11.9) | 57 (12.4) | 61.24 (11.01) |
| Systolic Blood Pressure, mm/Hg (Mean, SD) | 133.2 (14.8) | 130.3 (18.6) | 133.8 (17.9) | 132.1 (15.3) |
| BMI, kg/m<sup>2</sup>, (Mean, SD) | 27.7 (5.0) | 32.5 (7.5) | 26.8 (5.6) | 26.6 (5.5) |
| Haemoglobin, g/L (Mean, SD) | 13.3 (1.4) | 13.8 (1.6) | 13.1 (1.7) | 13.2 (1.9) |
| HbA1C, % (Mean, SD) | 8.6 (1.8) | 7.6 (1.9) | 8.0 (1.8) | 7.4 (1.6) |
| FBG, mmol/L., (Mean, SD) | 8.9 (3.2) | 8.8 (3.6) | 8.7 (3.2) | 8.1 (3.3) |
| Cholesterol, mmol/L., (Mean, SD) | 4.4 (1.1) | 4.6 (1.1) | 4.5 (1.0) | 4.4 (1.2) |
| HDL, mmol/L., (Mean, SD) | 1.1 (0.3) | 1.3 (0.4) | 1.2 (0.4) | 1.4 (0.5) |
| LDL, mmol/L., (Mean, SD) | 2.5 (0.8) | 2.6 (0.9) | 2.6 (0.8) | 2.5 (1.0) |
| TG, mmol/L., (Mean, SD) | 1.8 (2.2) | 1.6 (0.8) | 1.5 (1.0) | 1.9 (1.4) |
| Serum Creatinine, umol/L., (Mean, SD) | 74.2 (26.9) | 85.0 (56.3) | 74.0 (22.7) | 70.8 (23.3) |
| ACR, mg/mmol, (Mean, SD) | .. | .. | 1.3 (0.8) | .. |
| Serum Albumin, g/L (Mean, SD) | .. | .. | .. | 40.1 (5.3) |
| ALT, U/L (Mean, SD) | .. | .. | .. | 33.4 (44.5) |
| AST, U/L (Mean, SD) | .. | .. | .. | 34.2 (45.7) |
| Haematocrit, %, (Mean, SD) | .. | .. | .. | 39.6 (5.2) |

**Table 2.** List of parameters used for GMF in the identification model (EVAS and NHANES), incomplete input prediction model (CDMD) and complete input prediction model (SDR).

| **Parameters[Units]** | **EVAS** | **NHANES** | **CDMD** | **SDR** |
| --- | --- | --- | --- | --- |
| Fasting Blood Glucose (FBG) [mmol/L] | ✓ | ✓ | ✓ | ✓ |
| Total Cholesterol [mmol/L] | ✓ | ✓ | ✓ | ✓ |
| High density lipoprotein (HDL) [mmol/L] | ✓ | ✓ | ✓ | ✓ |
| Low density lipoprotein (LDL) [mmol/L] | ✓ | ✓ | ✓ | ✓ |
| Triglyceride [mmol/L] | ✓ | ✓ | ✓ | ✓ |
| Serum Creatinine [μmol/L] | ✓ | ✓ | ✓ | ✓ |
| Body Mass Index (BMI) [kg/m<sup>2</sup>] | ✓ | ✓ | ✓ | ✓ |
| HbA1c [%] | ✓ | ✓ | ✓ | ✓ |
| Haemoglobin (Hb) [g/dL] | ✓ | ✓ | ✓ | ✓ |
| Systolic Blood Pressure (SBP) [mm/Hg] | ✓ | ✓ | ✓ | ✓ |
| Albumin to Creatinine Ratio (ACR) [mg/mmol] | X | X | ✓ | X |
| Serum Albumin [g/L] | X | X | X | ✓ |
| Alanine Aminotransferease (ALT) [U/L] | X | X | X | ✓ |
| Aspartate Aminotransferase (AST) [U/L] | X | X | X | ✓ |
| Haematocrit [%] | X | X | X | ✓ |

### 2.2 GMF digital twins for the identification and prediction of CKD

To determine the accuracy and robustness of our generalized metabolic flux (GMF) based digital twin model, we first sought to investigate its performance in identifying CKD, predicting CKD and finally to stratify patients based on CKD risk (Figure 1). For the identification of CKD, our model achieved an AUC of 0.80 in the EVAS data set and an AUC of 0.82 in the NHANES data set (Figure 2a and b). To confirm the robustness of our model, we also performed another CKD identification analysis with a reduced set of input parameters and achieved reasonable performance (AUC 0.75 in EVAS and 0.73 in NHANES) (Supplementary Fig. 2). For the prediction of CKD within a 3 year time horizon, we achieved AUCs between 0.75 and 0.86 (Figure 2c and d). With incomplete parameters in the CDMD data set, we achieved an AUC of 0.75, whereas with complete parameters in the SDR data set, we achieved an AUC of 0.86. The full performance metrics with complete parameters in the SDR data set (AUC, Sensitivity (SN), Specificity (SP), Negative Predictive Value (NPV), Positive Predictive Value (PPV)) is shown below (Table 3). We achieved the same AUC values in both the testing sets, Testing-1 and Testing-2.

**Table 3.** Performance metrics achieved in the complete inputs prediction model analysis of the SDR data set.

| Metrics | Testing-1 | Testing-2 |
| --- | --- | --- |
| AUC | 0.86 | 0.86 |
| SN | 80% | 80% |
| SP | 72% | 74% |
| NPV | 84% | 85% |
| PPV | 65% | 66% |

**Fig. 1:**
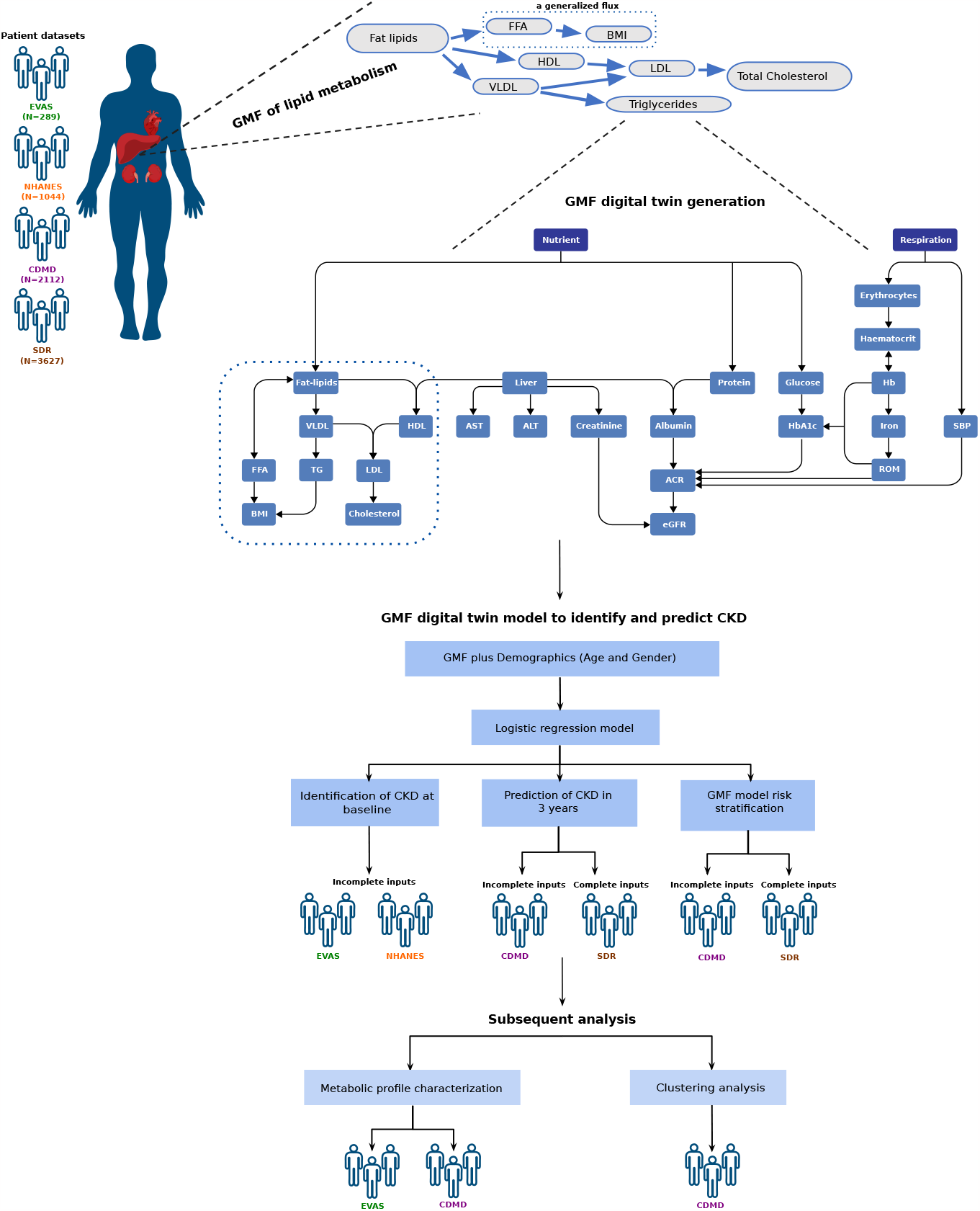
Study design. The scheme illustrates the data sets employed and how the generalized metabolic fluxes (GMF) digital twins model was created and subsequently analyzed in this study. From known biological relationships, the GMF processes are constructed to form the GMF digital twin model. The performance and capabilities of this model were tested in three main analyses, namely in the identification and prediction of chronic kidney disease (CKD), metabolic profile characterization and finally the clustering analysis.

**Fig. 2:**
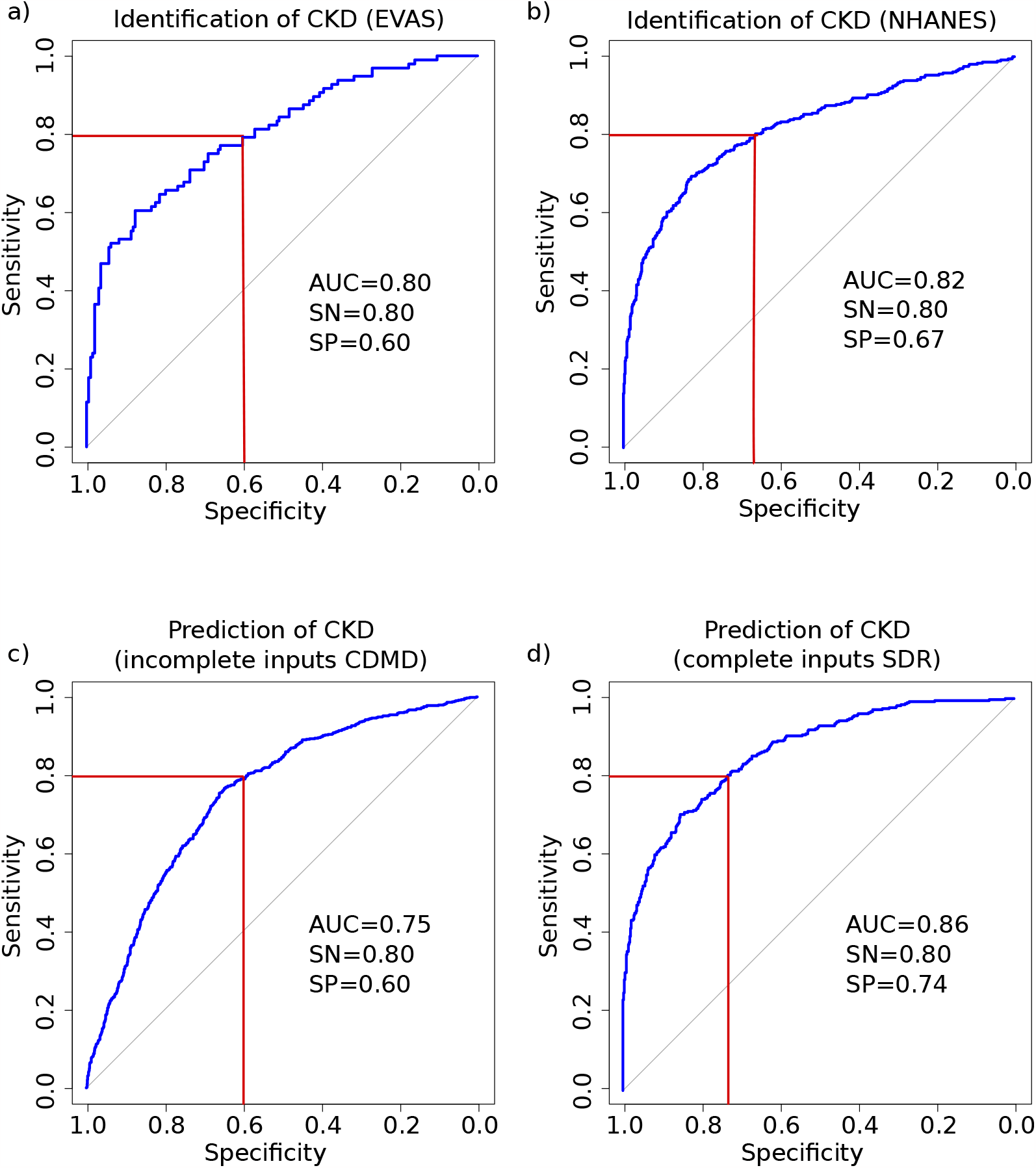
AUC-ROC curves for identification of chronic kidney disease (CKD) at baseline and the prediction of CKD within 3 years. The identification of CKD yielded an AUC of 0.80 in the EVAS (a) and 0.82 in the NHANES (b) data sets. The prediction of CKD yielded an AUC of 0.75 in the CDMD (c) and 0.86 in the SDR (d) data sets.The linear regression models for AUC-ROC curve generations utilized the GMF model, age and gender as input parameters. The SN values represent the Sensitivity, whereas the SP values represent the Specificity for each of the four AUC-ROC analyses.

After establishing the predictive capabilities of our model, we wanted to determine if it was also capable of stratifying patients into three different risk groups (high, moderate, low) accurately. The distribution of patients across the three risk groups (CDMD and SDR datasets) are presented in Table 4.. Most patients who developed CKD in 3 years came from the high risk group. In the CDMD data set, 53.3% of patients developed CKD in the high-risk group, 17.3% developed CKD in the moderate-risk group and 9.8% developed CKD in the low-risk group. Whereas in the SDR data set, 62.9% of patients developed CKD in the high-risk group, 19.3% developed CKD in the moderate-risk group and 5.4% developed CKD in the low-risk group. The observed frequencies in both data sets align with the expected risk frequency intervals reported in previous studies. [7, 8].

**Table 4.** Risk group classification using GMF with the incomplete inputs prediction model (CDMD) and the complete inputs prediction model (SDR)

| Risk group | CDMD |  |  | SDR |  |  | Risk interval |
| --- | --- | --- | --- | --- | --- | --- | --- |
|  | no CKD | CKD | % CKD | no CKD | CKD | % CKD | Expected CKD % |
| **High (>30%)** | 461 | 526 | 53.3 | 174 | 295 | 62.9 | 65% (30-100%) |
| **Moderate (10-30%)** | 886 | 188 | 17.3 | 205 | 49 | 19.3 | 20% (10-30%) |
| **Low (<10%)** | 46 | 5 | 9.8 | 174 | 10 | 5.4 | 5% (0-10%) |

### 2.3 Characteristics of patient GMF profiles and their association with future CKD development

We first looked at the correlation between the input parameters (clinical and physiological) used to create GMF with the GMF values itself using the CDMD data set. The input parameters show strong correlation with the relevant GMFs (Figure 3). Serum creatinine which is an indicator for CKD, has a strong positive correlation with individual GMF fluxes related to the circulation-respiratory pathways, the reactive oxygen species and HbA1c production pathways and negative correlation to the albumin-ACR pathways. Furthermore, the parameters LDL, Cholesterol, BMI, HbA1c and Glucose seem to cluster together, where LDL, Cholesterol and BMI also show strong positive correlation to the GMF fluxes related to lipid metabolism. To determine factors that could predict future categorization into CKD positive or negative groups, we studied the baseline GMF profiles used for this prediction. We conducted two subgroup analyses: one comparing metabolic profiles of future CKD (CDMD) and future non-CKD patients (CDMD), and another comparing baseline CKD (EVAS) and future CKD patients (CDMD) (Figure 4). Future CKD patients exhibited elevated fluxes associated with circulation, blood pressure, glucose metabolism, and kidney function (Figure 4, Supplementary Table 4). This clearly shows that future CKD patients exhibit a more deteriorated health state (GMF profile) compared to future non-CKD patients. When then compared patients who develop CKD in the future with patients with CKD at baseline. Patients who develop CKD in the future displayed decreased fluxes in glucose and lipid metabolism pathways, as well as pathways related to kidney function, respiration, and circulation, compared to patients who had CKD at baseline, (Figure 4, Supplementary Table 4). As expected, the health state (GMF profile) of future CKD patients were less deteriorated compared to baseline CKD patients.

**Fig. 3:**
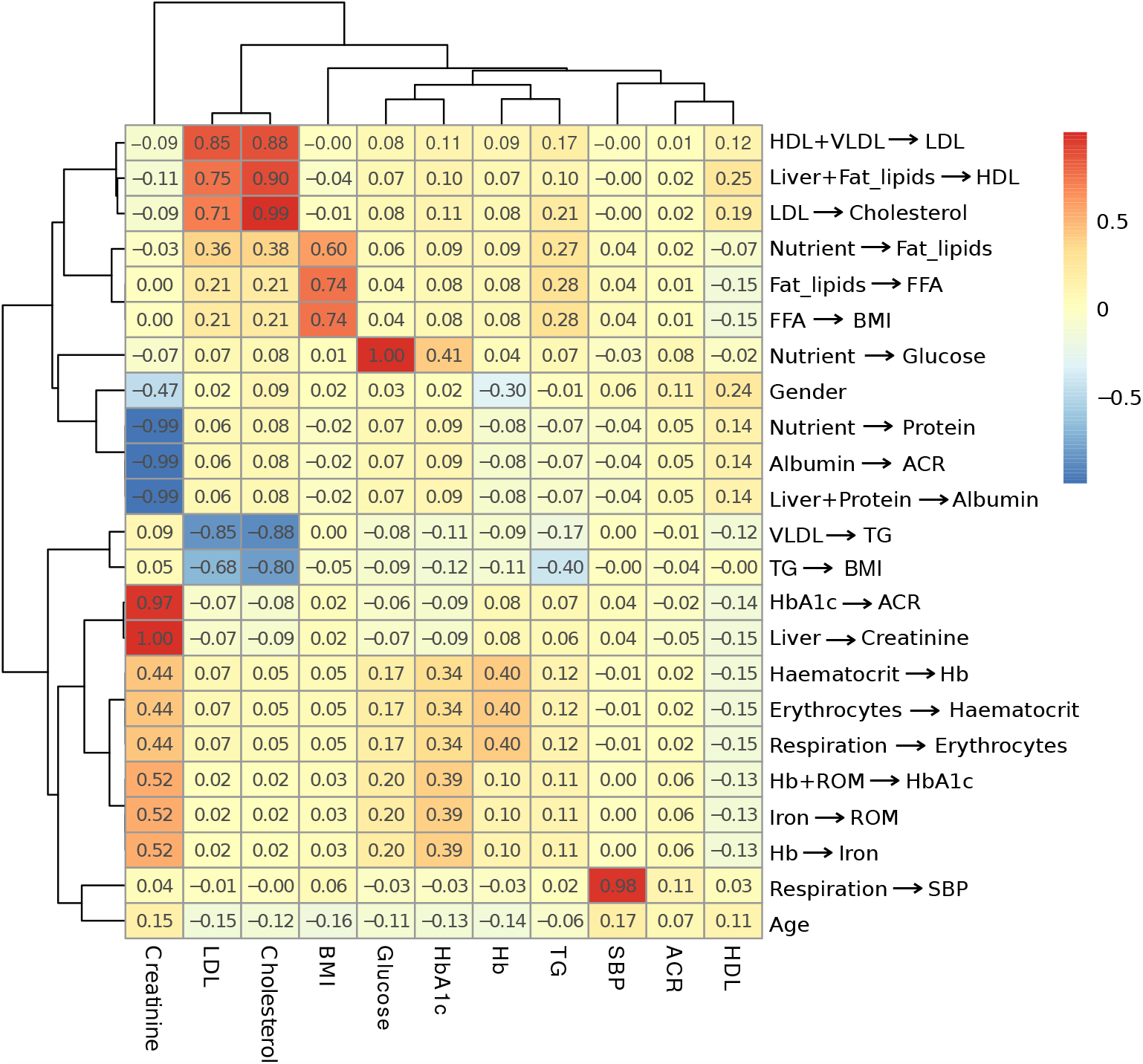
Correlation between input parameters and GMF. The correlation between clinical and physiological input parameters with GMF is shown here. The numbers in each box represent the Kendall’s *τ* correlation value between two variables, the inputs vs the GMFs. Serum creatinine is correlated with a number of individual GMFs, particularly the GMFs related to the respiration-circulation pathways, reactive oxygen species and HbA1c production pathways and the albumin-ACR pathways. LDL, Cholesterol, BMI, glucose and HbA1c cluster together, whereby LDL, Cholesterol and BMI are strongly correlated to the GMFs related to the lipid metabolism pathways.

**Fig. 4:**
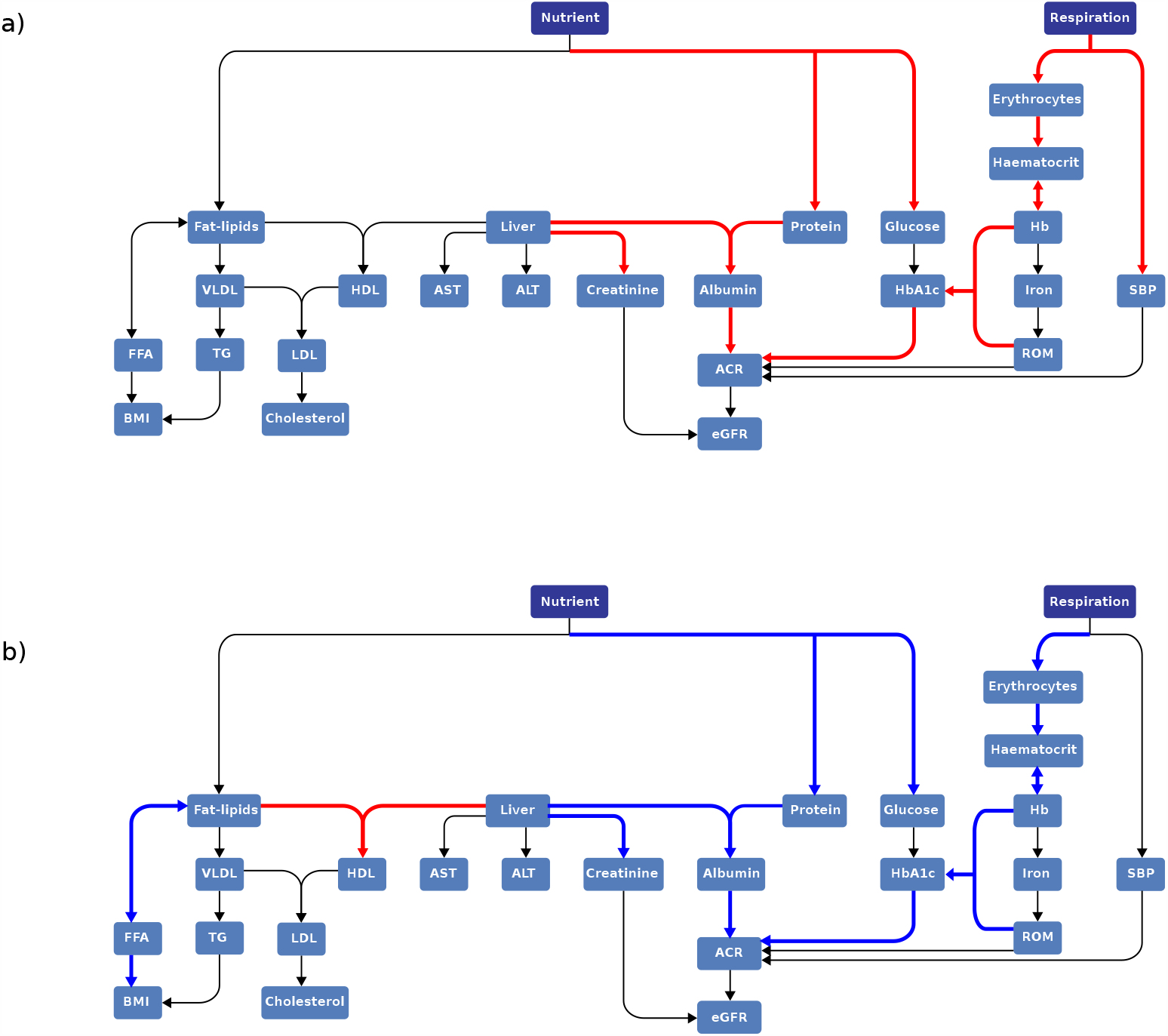
Metabolic profile characteristics in future non-CKD vs future CKD and baseline CKD vs future CKD. Two GMF profiles from 2 subgroup analysis of patients are shown here. The GMF profile of patients who develop CKD in the future shows a poorer health profile than the GMF profile of patients who do not develop CKD in the future (a). The GMF profile of patients who develop CKD in the future shows a better health profile than the GMF profile of patients who have CKD at baseline (b).

### 2.4 Distance between patient profile clusters indicates the rate of future CKD development

We found that among CKD-negative patients at baseline, some profiles resembled CKD patients while others did not. Thus, we sought a metric to distinguish these profiles and predict CKD development. We conducted patient clustering in the CDMD data set using two sets of variables: the first set consisted of GMF values, while the second set utilized input parameters. The GMF-based patient cluster with the highest fraction of future CKD outcomes was the most distinct cluster from all other clusters (Figure 5a). In this cluster, the fluxes related to circulation-respiratory pathways as well as the reactive oxygen, creatinine and HbA1c production pathways were increased whereas the fluxes related to the albumin-ACR pathways were decreased. To analyze the relationship between GMF profiles and CKD development, we ordered the clusters by increasing CKD outcome rates. We picked the cluster with the lowest fraction of positive CKD outcomes as the starting point. For all other clusters, we computed the Euclidean distance from their centers to the starting point. We examined the dependence of the CKD outcomes rate in each cluster on this distance metric. There is a significant positive correlation (*τ* = 0.6, p = 0.017) between the distance metric and CKD outcome rates when we used GMF to cluster patients (Figure 5b). The cluster with the highest fraction of positive CKD outcomes was the furthest away from the cluster with the lowest fraction of positive CKD outcomes. By contrast, we found no distinct cluster profiles and no significant associations between the distance metric and CKD outcome rates when we used input parameters for clustering (Supplementary Fig. 4).

**Fig. 5:**
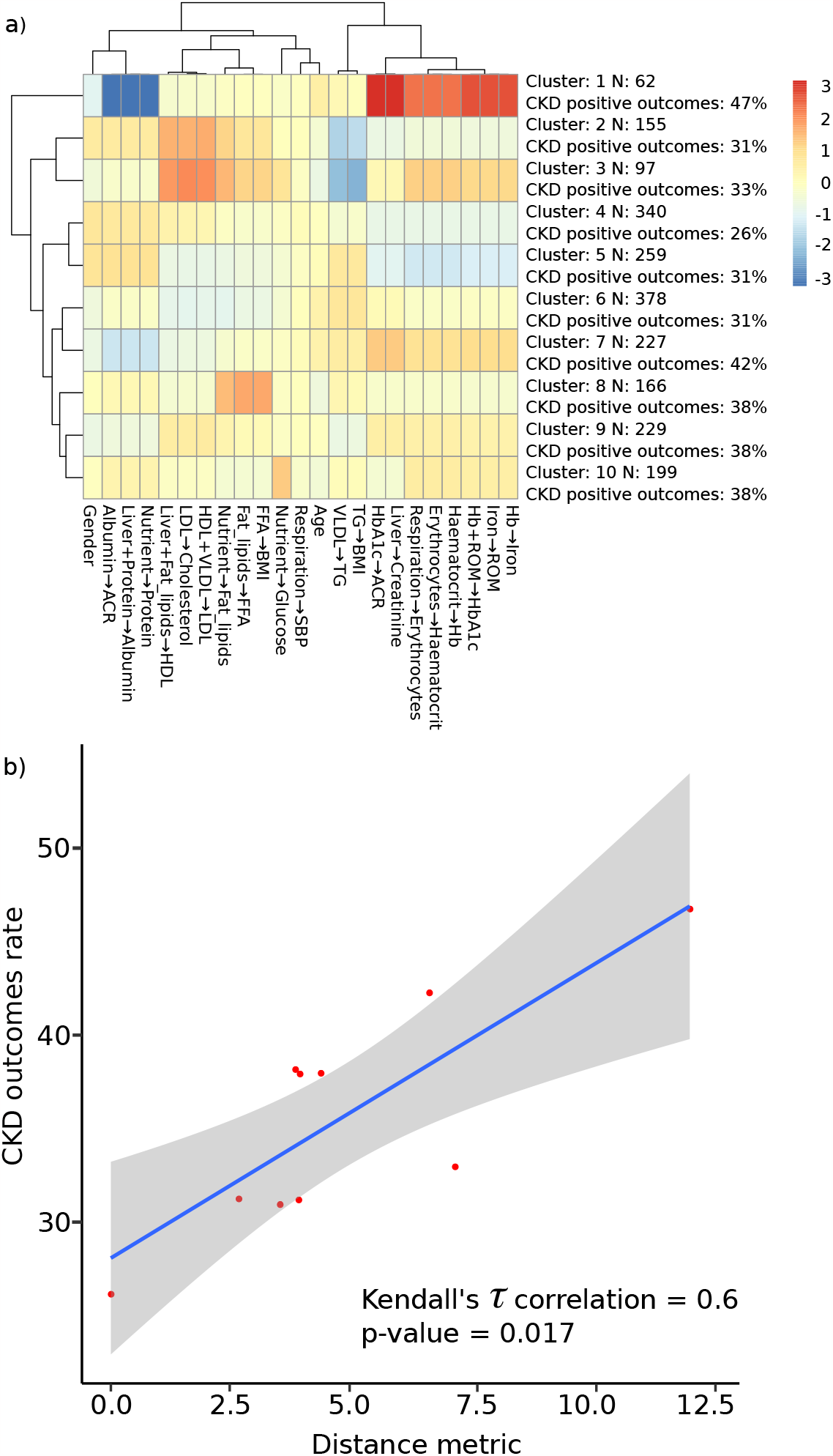
Patient clustering with GMF. The figure illustrates the clustering pattern of patients in the CDMD dataset using GMF (a) and the relationship between the cluster distances and CKD outcomes rate (b). N represents the number of patients within the specific cluster, and CKD positive outcomes are calculated as the ratio of patients developing future CKD (within 3 years) to the total number of patients in that cluster. The cluster with the highest CKD outcomes rate has a distinct pattern with elevated flux values related to the respiration-circulation pathways, reactive oxygen species and HbA1c production pathways and the albumin-ACR pathways. There is a significant correlation (*τ*=0.6, p=0.017) between cluster distance and CKD outcomes rate where the cluster with the highest CKD outcomes rate is furthest away from the cluster with the lowest CKD outcomes rate. Standard error (SE) is denoted by the grey shaded area.

## 3 Discussion

Here, we developed a generalized metabolic fluxes (GMF) based digital twin model called HealthVector Diabetes (HVD) [6] to identify CKD and to predict the occurrence of CKD within 3 years in T2DM patients. The HVD GMF digital twin model describes the current health state of a patient and predicts their future health trajectory. In retrospective clinical data sets, our model achieved a high performance (AUC=0.8-0.82) in describing CKD patients by their metabolic states at baseline. Remarkably, our model also performed well at predicting CKD within a 3 year time horizon (AUC 0.75-0.86) in two scenarios, with imcomplete and complete parameters.

Our model identified metabolic health profile differences at baseline showing that patients who develop future CKD have more deteriorated health profiles compared to patients who do not. Our model also stratified the patients into high, moderate, and low-risk groups. The highest percentage of patients who developed CKD was found in the high-risk group (53.9-62.9%), followed by the moderate-risk group (17.3-19.3%), and finally the low-risk group (5.4-10.7%). These results lay the groundwork for future clinical applications of this model for CKD risk assessment and personalized care planning in T2DM populations.

There are several machine learning models that have been developed for detection or prediction of CKD. One detection model using simple screening parameters (age, gender, BMI, waist circumference, and urine dipstick) achieved moderate performance (AUC = 0.76) [9]. Another detection model detected CKD from retinal images (AUC=0.93), using a deep learning algorithm [10]. Despite the high performance of this model, retinal imaging introduces added costs and time, thereby diminishing the feasibility of this procedure. In contrast, our model utilizes readily available parameters obtained from routine patient blood screening (Table 1), while achieving AUCs of 0.80-0.82. Our model is better at CKD prediction than most published models that use single time point readings (biochemical or otherwise) as their inputs [8, 11–15]. Other published models have lower AUC values and they use non-routine measurements as inputs (Supplementary Table 5). Compared to these models, our model was evaluated in diverse cross-country multi-center and multi-ethnic cohorts emphasizing its applicability.

Our incomplete parameter prediction model using the CDMD data set utilized 11 common input parameters to construct the GMF digital twins. Given the inherent characteristics of this EMR data set, a significant proportion of missing data had to be addressed. Strictly adhering to complete data points would have substantially reduced our sample size. To strike a balance, we allowed for data points with up to one missing parameter. Despite the presence of missing parameters, this model exhibited reasonable performance with an AUC of 0.75. Subsequently, in another data set (SDR), we exclusively worked with complete data points and explored the inclusion of additional input parameters. Comparatively, our complete parameter prediction model employed 14 common inputs and demonstrated superior performance (AUC=0.86). Notably, complete data points are not always attainable in clinical settings, and most models struggle to handle missing data. Our GMF model demonstrated good performance in both the complete and the incomplete input data scenarios, validating their suitability for real world clinical applications. Currently, routine clinical follow-ups fail to detect patients at risk of developing CKD due to their kidney health indicators (ACR or eGFR) typically falling within normal ranges and below diagnostic thresholds. In a clinical context, understanding the metabolic GMF profiles of at-risk patients would guide clinical decisions aimed at preventing future disease states or potentially reversing current disease states. Leveraging our GMF model, clinicians can identify and stratify T2DM patients before CKD manifestation, enabling targeted personalized care for the patients at risk.

The GMF digital twin model informs the long-term rates of metabolic changes and predict the evolution of metabolic characteristics of CKD [6]. The model can identify early indicators of health deterioration to predict future health states by highlighting key pathways involved in dysfunctions leading to kidney failure. Our analysis reveals that patients who will develop CKD in the future exhibit an overall deteriorated metabolic profile, as indicated by elevated individual GMF or metabolic rates for fluxes related to glucose production and kidney function compared to patients who do not develop CKD (Figure 4, Supplementary Table 4). Furthermore, our model effectively differentiates between the health states of patients with CKD at baseline and those who develop CKD in the future. Patients who only develop future CKD have a considerably healthier metabolic profile as indicated by reduced metabolic rates for fluxes related to glucose production and kidney function compared to patients already diagnosed with CKD (Figure 4, Supplementary Table 4). Consistent with the results of our predictive analysis, patients with CKD at baseline exhibited more deteriorated health profiles compared to patients without CKD at baseline (Supplementary Fig. 3).

GMF profiles are also able to cluster patients according to their metabolic characteristics which in turn reflect the degree of health deterioration. These metabolic changes in the GMF profiles drive the increased rate of future CKD outcomes. The fluxes related to respiratory-circulation, glucose metabolism and kidney function were distinctly elevated in the cluster with the highest CKD outcome rates. These fluxes overlap with those identified in the previous analysis where we compared the GMF profiles of future CKD and future non-CKD patients. The partial overlap is expected due to cluster heterogeneity, encompassing both future CKD and future non-CKD patients in every cluster. Measuring the distance between cluster centers reveals a positive correlation between cluster distance and CKD outcome rates. The cluster with the highest positive CKD outcomes is farthest from the cluster with the lowest positive CKD outcomes. This highlights GMF’s ability to map baseline health states to patient health trajectories. While these clusters encompass a mixture of patients, including patients who develop future CKD and those who do not, it is important to emphasize that our study focuses on a 3-year time horizon. Future investigations considering a more extended time frame may reveal an increased fraction of positive CKD outcomes in the clusters characterized by greater health deterioration. Basic clinical and physiological input parameters failed to demonstrate distinct patient cluster profiles and exhibited no correlation with cluster distances and CKD outcome rates (Supplementary Fig. 4).

While our study has demonstrated promising results, it is important to acknowledge the limitations that exist. Firstly, in the CDMD and SDR real world data sets, we applied data aggregation to obtain the input parameter values. This involved taking the median values of each parameter with repeated measurements over a single year. Since the measurements for each parameter were not obtained on a single day, this led to less stringent control over the variables and introduced non-uniformity, particularly in parameters such as ACR (Supplementary Fig. 1). It is also well known that ACR measurements can be highly variable in individuals [10]. Consequently, ACR was not included in our final prediction model (SDR) (Table 2). We also did not consider the impact of medications on patients health status and how this would impact our GMF model. We know that treatment interventions can significantly influence patient’s health states. Therefore, utilizing our model to assess the effect of medication on patient’s metabolic health profiles could offer valuable insights. These limitations highlight areas for future research and improvement in our approach.

In conclusion, we have developed a digital twin GMF model named HealthVector Diabetes (HVD) that successfully identifies existing metabolic pathways dysfunctional in CKD patients, and predicts future CKD within a 3 year time horizon. Our model has great potential to be adopted in the clinical setting as a predictive tool in the prevention of CKD amongst T2DM patients.

## 4 Methods

### 4.1 Study design and data set characteristics

Our study included four populations: three multi-ethnic T2DM cohorts from Singapore and one multi-ethnic T2DM North American cohort. Our first dataset (EVAS) was a multi ethnic cohort of T2DM patients from Singapore’s Tan Tock Seng Hospital (TTSH) that were part of a clinical study and were followed-up for 5 years between 2015 to 2020 [16]. The second data set was obtained by selecting T2DM patients from data collected in the National Health and Nutrition Examination Survey (NHANES) that were recruited between 1999 to 2018 (NHANES dataset) [17]. The third dataset (CDMD), the National Healthcare Group Chronic Disease Management Datamart (NHG-CDMD) was obtained from the electronic medical records (EMR) of TTSH, Singapore between 2008 to 2021. The fourth data set was a group of T2DM patients extracted from a health database registry of Singapore Health Services (SingHealth), the SingHealth Diabetes Registry (SDR) between 2013 to 2020 [18]. All patients used in the four data sets were aged between 20 to 80 years and their baseline characteristics are outlined below (Table 1). Baseline individual generalized metabolic flux (GMF) profiles were calculated for each individual from the selected list of clinical and physiological parameters relevant to CKD (Table 2). We conducted the following analyses: i) identification of CKD in patients ii) prediction of future CKD within three years with risk stratification, iii) metabolic profile characterization and correlation, and iv) patient clustering with either GMF or input parameters. For the identification of CKD, we used the EVAS and NHANES data sets and for the prediction of CKD and risk stratification, we used the CDMD and SDR data sets. Subsequently, for the metabolic profile characterization analysis, we used the CDMD and EVAS data sets and finally for the correlation and clustering analysis, we only used the CDMD data set. Ethics approval was obtained from the Singaporean (NHG and SingHealth) Institutional Review Board. This study was conducted in accordance to the principles of the Declaration of Helsinki.

### 4.2 Generalized metabolic fluxes (GMF) digital twin generation and model development

In this study, we utilized Generalized Metabolic Flux (GMF) models to create personalized digital twins for each patient [6]. GMF models comprise a network of dynamic variables, which represent the metabolic rates of the patient at a specific health state. We distinguished two reference health states: A) T2DM without CKD and B) T2DM with CKD. At the baseline time point, patients were either in state A or state B and using GMF, we described their health state at that time point. On the other hand, during the course of our prediction study, some patients progressed from state A to state B within a defined period of time (i.e., 3 years), while others remained in state A. For each patient, the progression occurred along a single health state progression scale, which we termed a generalized extent, spanning between the two basic states: A and B. This scale depicts a continuous change in the patient’s metabolic profile. As a dynamic variable, a single GMF measures the rate of change of a specific metabolite concentration or a physiological reading at any given state along the progression scale. The collection of individual GMFs for a particular patient at a particular time point represents that patient’s metabolic digital twin. The GMF methodology allows us to use both complete and incomplete inputs to produce digital twins of patients and best fit models. We investigated the performance of each (incomplete and complete) in the two separate data sets (CDMD and SDR) for the prediction of CKD. The completeness of inputs in each data set is characterized in the Supplementary Materials (Supplementary Table 2). These GMFs combined make up the GMF profile for each patient in each studied cohort and represent the patient’s digital twin (Figure 1, Supplementary Table 3). The detailed method of producing patients GMF digital twins has been explained in our earlier technical paper [6]. HealthVector Diabetes (HVD) is an identification and prediction model based on GMF digital twins.

For the identification of CKD, the GMF model employed 10 clinical and physiological parameters as inputs and produced 21 informative GMF fluxes as outputs. The outputs were combined with the patients’ age and gender information to build the logistic regression model for identification of CKD cases in the EVAS and NHANES datasets. Similarly, for the prediction of CKD, there were two models: i) the model with incomplete parameters and ii) the model with complete parameters. For the incomplete parameters, the GMF model employed 11 parameters as inputs whereas for the complete parameters, the GMF model employed 14 parameters as inputs. The incomplete parameter model was tested in the CDMD data set whereas the complete parameter model was tested in the SDR data set. The comprehensive list of input parameters utilized in the analysis of each data set is shown below (Table 2).

### 4.3 Clinical definitions and parameter selection

Clinical measurements were taken at their point of recruitment (all data sets) and follow-up (CDMD and SDR). In the EVAS, NHANES, and CDMD data sets, patients were identified to have CKD if they fell under one of the following two categories: i) if they had an Albumin to Creatinine Ratio (ACR) value of more than 3.3 mg/mmol (equivalent to 30 mg/g) ii) if they had an estimated glomerular filtration rate (eGFR) value of less than 60 mL/min/1.73*m*^2^ [19]. For the SDR data set, we used only the second criterion: the eGFR value of less than 60 mL/min/1.73*m*^2^. This was due to the SDR data set being primarily composed of primary care patients whereas the EVAS and CDMD data set were primarily composed of tertiary care patients (Supplementary Fig. 1). For the EVAS and CDMD data sets, the eGFR value was calculated using the New Asian Modified CKD-EPI formula because this was the equation used by the clinician in charge of these data sets in TTSH. [20] Whereas for the NHANES and the SDR data sets, the standard Chronic Kidney Disease Epidemiology Collaboration (CKD-EPI) formula was used. [21]

### 4.4 Logistic regression and risk stratification analysis

We first performed the analysis on the identification of CKD disease state at baseline to determine the present health state of patients using two data sets, the EVAS and the NHANES data sets. Subsequently, we predicted the CKD disease state within three years using two separate models, the incomplete parameter model in the CDMD data set and the complete parameter model in the SDR data set to assess the performance of our model in predicting the health state of patients within 3 years. To this end, we developed a logistic regression (LR) model using GMFs derived from basic parameter inputs (Table 2), age, and gender as predictor variables, where we plotted the receiver operating characteristic (ROC) curve and estimated the area under the curve (AUC). In the complete parameter prediction model, the SDR data set was randomly split into the training set (50% of the population) and two testing sets, Testing-1 (25%) and Testing-2 (25%). We trained the model on the training set to obtain the parameters of the LR model, which was then used to predict the probability of CKD in the Testing-1 and Testing-2 sets.

To evaluate the efficacy of our model in stratifying patients into high-risk, moderate-risk, and low-risk groups for CKD, we compared the observed frequency of future CKD positive outcomes in each risk group to the expected frequency of CKD positive outcomes investigated in the CDMD and SDR data sets. The expected frequency of the risk groups was based on previously published risk intervals [7, 8]. High risk is classified as having 30-100% CKD patients, moderate risk as having 10-30% CKD patients and low risk as having 0-10% CKD patients.

### 4.5 Metabolic profile characterization, correlation and clustering analysis

We examined the correlation between the input parameters, GMFs, and demographic values. Kendall’s *τ* correlation method was employed for this assessment. In cases of missing values, we substituted them with the median values from the CDMD dataset. To identify groups of correlated parameters, we utilized hierarchical clustering, using the Euclidean distance metric and the complete linkage method on the resultant correlation matrix (Figure 3).

We then performed a metabolic profile subgroup analysis to explain the variation in the GMF profiles in different groups of patients, based on inputs at baseline. First, we investigated the GMF differences seen in patients who develop CKD in the future with respect to patients who do not develop CKD in the future. Second, we looked at the GMF profile differences in patients who develop CKD in the future with respect to patients with CKD at baseline. The grouped median values of each individual GMF within the entire GMF profile were compared in each of the subgroup analyses. The significant difference of GMF values between the groups were evaluated using the Wilcoxon-Mann-Whitney U test with the two-sided null hypothesis (Supplementary Table 4). The GMF profiles are visualized in a standard graphical form, wherein each individual GMF within the entire GMF profile is represented as colored directed edges (Figure 4). The color on the map corresponds to the ratio between the median value of each individual GMF in a specific group and its median in the reference group. Elevated and reduced GMFs that were significant in one subgroup vs the other subgroup are shown in red and blue, respectively. The GMFs that do not vary are shown in black.

We next investigated the relationship between future CKD outcomes and clusters of patients. To reveal associations between the groups of patients and their metabolic features, we applied k-means clustering. To determine the optimal number of clusters (k), we assessed the Between Sum of Squares (BSS) metric over a range of k values from 5 to 20. The BSS with 50% was achieved with k = 10 and this k was selected as the optimal choice for our analysis. The clusters were assigned numerical indices and ordered in ascending order of CKD outcome rates. The cluster with the lowest fraction of future CKD-positive patients was designated as index 1. For all indices, we computed the Euclidean distance between the centroid of the i-th index and the centroid of the cluster with index 1. These distances were then plotted against the fraction of future CKD positives in each cluster, and their correlation was evaluated using Kendall’s *τ* with the two-sided null hypothesis that no correlation is present.

### 4.6 Statistical analysis and computational tools

All statistical analyses were performed using R version 3.6.3. For logistic regression analysis, we utilized the glm.fit function within the core stats package in R [22]. The ROC curves were generated using the pROC R package [23, 24]. The Wilcoxon-Mann-Whitney U test from the core stats package in R was used to determine the significant difference of the grouped median fluxes in the different patient subgroups [23]. For the correlation analysis, the cor function was used within the core stats package [23]. Subsequently, the pheatmap package was used to visualize correlation outputs [25]. The pheatmap package was also used to cluster patients with k-means and to visualize the clusters [23, 25].

## Data Availability

The authors agree to make the data and materials supporting the results or analyses presented in their paper available upon reasonable request. Access to the Singapore Diabetes Registry data set or the other data sets is possible on reasonable request to the corresponding authors, under restrictions subject to obtaining ethics approval from institutional boards and an appropriate data-use and/or research agreement.

## Declarations

### 4.8 CODE AVAILABILITY

The code in this study is available from the authors upon reasonable request.

## 4.9 Acknowledgements

We would like to acknowledge Prof. Frank Eisenhaber for insights and fruitful discussions on the biological and mathematical aspects of the work. We would like to acknowledge Dr. Jonathan Wei Xiong Ng for his exploratory analysis and insights that increased the clarity of the scope of the study. The work was supported by Enterprise SG, under the Startup SGTech POC and POV grant and also by A*STAR, Singapore under its Industry Alignment Pre-Positioning Fund (Grant No. H19/01/a0/023 – Diabetes Clinic of the Future). RD is supported by Ministry of Health, Clinician Scientist Award (MOH-000014), Ng Teng Fong Foundation Grant and National Healthcare Group. The funding bodies played no role in the design of the study and collection, analysis, interpretation of data or in writing the manuscript.

## 4.10 Author contributions

This study was designed and planned by AB, NUS, RD, and YMB. NUS and AB analyzed the data. The clinical data sets were provided by RD, SY and YMB along with clinical and methodological expertise. AW and WL provided strategic guidance and oversight. NUS and AB drafted the manuscript. Writing, review and editing were performed by all authors. The final version of the paper has been seen and approved by all the authors. Funding acquisition for this study was by RD, AW, AB and WL.

## References

[1] Sikdar KC, Wang PP, MacDonald D, Gadag VG. Diabetes and its impact on health-related quality of life: a life table analysis. Quality of Life Research. 2010 mar;19(6):781–787. 10.1007/s11136-010-9641-5.

[2] Bee YM, Tai ES, Wong TY. Singapore’s “War on Diabetes”. The Lancet Diabetes and Endocrinology. 2022 jun;10(6):391–392. 10.1016/s2213-8587(22)00133-4.

[3] Chen HY, Kuo S, Su PF, Wu JS, Ou HT. Health Care Costs Associated With Macrovascular, Microvascular, and Metabolic Complications of Type 2 Diabetes Across Time: Estimates From a Population-Based Cohort of More Than 0.8 Million Individuals With Up to 15 Years of Follow-up. Diabetes Care. 2020 may;43(8):1732–1740. 10.2337/dc20-0072.

[4] Bruynseels K, de Sio FS, van den Hoven J. Digital Twins in Health Care: Ethical Implications of an Emerging Engineering Paradigm. Frontiers in Genetics. 2018 feb;9. 10.3389/fgene.2018.00031.

[5] Venkatesh KP, Raza MM, Kvedar JC. Health digital twins as tools for precision medicine: Considerations for computation, implementation, and regulation. npj Digital Medicine. 2022 sep;5(1). 10.1038/s41746-022-00694-7.

[6] Batagov A, Dalan R, Wu A, Lai W, Tan CS, Eisenhaber F. Generalized metabolic flux analysis framework provides mechanism-based predictions of ophthalmic complications in type 2 diabetes patients. Health Information Science and Systems. 2023 mar;11(1). 10.1007/s13755-023-00218-x.

[7] Jiang W, Wang J, Shen X, Lu W, Wang Y, Li W, et al. Establishment and Validation of a Risk Prediction Model for Early Diabetic Kidney Disease Based on a Systematic Review and Meta-Analysis of 20 Cohorts. Diabetes Care. 2020 mar;43(4):925–933. 10.2337/dc19-1897.

[8] Chan L, Nadkarni GN, Fleming F, McCullough JR, Connolly P, Mosoyan G, et al. Derivation and validation of a machine learning risk score using biomarker and electronic patient data to predict progression of diabetic kidney disease. Diabetologia. 2021 apr;64(7):1504–1515. 10.1007/s00125-021-05444-0.

[9] Bradshaw C, Kondal D, Montez-Rath ME, Han J, Zheng Y, Shivashankar R, et al. Early detection of chronic kidney disease in low-income and middle-income countries: development and validation of a point-of-care screening strategy for India. BMJ Global Health. 2019 sep;4(5):e001644. 10.1136/bmjgh-2019-001644.

[10] Sabanayagam C, Xu D, Ting DSW, Nusinovici S, Banu R, Hamzah H, et al. A deep learning algorithm to detect chronic kidney disease from retinal photographs in community-based populations. The Lancet Digital Health. 2020 jun;2(6):e295–e302. 10.1016/s2589-7500(20)30063-7.

[11] Dong W, Wan EYF, Fong DYT, Kwok RLP, Chao DVK, Tan KCB, et al. Prediction models and nomograms for 10-year risk of end-stage renal disease in Chinese type 2 diabetes mellitus patients in primary care. Diabetes, Obesity and Metabolism. 2021 jan;23(4):897–909. 10.1111/dom.14292.

[12] Zhang K, Liu X, Xu J, Yuan J, Cai W, Chen T, et al. Deep-learning models for the detection and incidence prediction of chronic kidney disease and type 2 diabetes from retinal fundus images. Nature Biomedical Engineering. 2021 jun;5(6):533–545. 10.1038/s41551-021-00745-6.

[13] Lin CC, Niu MJ, Li CI, Liu CS, Lin CH, Yang SY, et al. Development and validation of a risk prediction model for chronic kidney disease among individuals with type 2 diabetes. Scientific Reports. 2022 mar;12(1). 10.1038/s41598-022-08284-z.

[14] Wu M, Lu J, Zhang L, Liu F, Chen S, Han Y, et al. A non-laboratory-based risk score for predicting diabetic kidney disease in Chinese patients with type 2 diabetes. Oncotarget. 2017 oct;8(60):102550–102558. 10.18632/oncotarget.21684.

[15] Sheng Qian Y, Moy FM. Predicting the Risk of Chronic Kidney Disease Among Type 2 Diabetes Mellitus Patients in a Primary Care Setting: An Evaluation of the QKidney Model. Malaysian Journal of Medicine and Health Sciences. 2019 10;15:67–73.

[16] Dalan R, Goh LL, Lim CJ, Seneviratna A, Liew H, Seow CJ, et al. Impact of Vitamin E supplementation on vascular function in haptoglobin genotype stratified diabetes patients (EVAS Trial): a randomised controlled trial. Nutrition and Diabetes. 2020 apr;10(1). 10.1038/s41387-020-0116-7.

[17] : Centers for Disease Control and Prevention. About the national health and nutrition examination survey. https://www.cdc.gov/nchs/nhanes/about_nhanes.htm.Accessed [11/11/2022].

[18] Lim DYZ, Chia SY, Kadir HA, Salim NNM, Bee YM. Establishment of the SingHealth Diabetes Registry. Clinical Epidemiology. 2021 mar;Volume 13:215–223.10.2147/clep.s300663.

[19] Goh S, Ang S, Bee Y, Chen Y, Gardner D, Ho E, et al. Ministry of Health Clinical Practice Guidelines: Diabetes Mellitus. Singapore Medical Journal. 2014 jun;55(6). 10.11622/smedj.2014079.

[20] Wang J, Xie P, Huang Jm, Qu Y, Zhang F, Wei Lg, et al. The new Asian modified CKD-EPI equation leads to more accurate GFR estimation in Chinese patients with CKD. International urology and nephrology. 2016;48(12):2077–2081. 10.1007/s11255-016-1386-9.

[21] Levey AS, Stevens LA, Schmid CH, Zhang YL, Castro AF, Feldman HI, et al. A New Equation to Estimate Glomerular Filtration Rate. Annals of Internal Medicine. 2009 may;150(9):604. 10.7326/0003-4819-150-9-200905050-00006.

[22] Müller M. Generalized linear models. In: Handbook of Computational Statistics. Springer; 2012. p. 681–709.

[23] R Core Team.: R: A Language and Environment for Statistical Computing. Vienna, Austria. ISBN 3-900051-07-0. Available from: http://www.R-project.org/.

[24] Robin X, Turck N, Hainard A, Tiberti N, Lisacek F, Sanchez JC, et al. pROC: an opensource package for R and S+ to analyze and compare ROC curves. BMC Bioinformatics. 2011;12:77.

[25] Kolde R. pheatmap: Pretty Heatmaps; 2019. R package version 1.0.12. Available from:https://CRAN.R-project.org/package=pheatmap.

