## Supplementary Materials for "A digital twin model incorporating generalized metabolic fluxes to predict chronic kidney disease in type 2 diabetes mellitus"

September 21, 2023

#### Tables

Table 1: List of Abbreviations

| **Abbreviations** | **Explanations** |
|---|---|
| ACR | Albumin-Creatinine Ratio |
| ALT | Alanine aminotransferase |
| AST | Aspartate aminotransferase |
| AUC | Area under the curve |
| BMI | Body mass index |
| CKD | Chronic Kidney Disease |
| eGFR | Estimated glomerular filtration rate |
| EMR | Electronic medical record |
| GMF | Generalized Metabolic Fluxes |
| FFA | Free fatty acids |
| FPG | Fasting plasma glucose |
| Hb | Haemoglobin |
| HbA1c | glycated haemoglobin |
| HDL | High-density lipoprotein |
| LDL | Low-density lipoprotein |
| ROC | Receiver operating characteristic |
| ROM | Reactive oxygen material |
| SBP | Systolic blood pressure |
| SD | Standard deviation |
| SN | Sensitivity |
| SP | Specificifty |
| TG | Triglyceride |
| VLDL | Very-low-density lipoprotein |

Table 2: Completeness of the data sets at baseline

| | EVAS (n =289) | NHANES (n=1044) | CDMD (n=2112) | SDR (n=3276) |
|---|---|---|---|---|
| Number of patients with 0 missing values | 92 (31.8%) | 1022 (98%) | 1228 (58%) | 3627 (100%) |
| Number of patients with 1 missing value | 132 (45.7%) | 22 (2%) | 884 (42%) | - |
| Number of patients with 2 missing values | 58 (20.1%) | - | - | - |
| Number of patients with 3 or more missing values | 7 (2.4%) | - | - | - |

Table 3: Digital twin map metabolic pathway reference sources

| **Pathway** | **Reference Source** |
|---|---|
| Respiration→Erythrocytes | Red blood cell pH, the Bohr effect, and other oxygenation-linked phenomena in blood O2 and CO2 transport.[1] |
| Erythrocytes→Haematocrit | Red blood cell pH, the Bohr effect, and other oxygenation-linked phenomena in blood O2 and CO2 transport.[1] |
| Iron→ROM | A general map of iron metabolism and tissue-specific subnetworks.[2] |
| Haematocrit↔Hb | Diagnostic morphology: biophysical indicators for iron-driven inflammatory diseases.[3] |
| Hb→Iron | Ferroptosis: Role of lipid peroxidation, iron and ferritinophagy.[4] |
| HDL+VLDL→LDL | The metabolic pathways of high-density lipoprotein, low-density lipoprotein, and triglycerides: a current review.[5] |
| Liver+Fat-lipids→HDL | High Density Lipoproteins: Metabolism, Function, and Therapeutic Potential.[6] |
| LDL→Cholesterol | Dyslipidaemia.[7] |
| VLDL→TG | The Effects of Medications Used for the Management of Diabetes and Obesity on Postprandial Lipid Metabolism.[8] |
| Fat-lipids↔FFA | New methodologies for studying lipid synthesis and turnover: Looking backwards to enable moving forwards.[9] |
| Fat-lipids→VLDL | New methodologies for studying lipid synthesis and turnover: Looking backwards to enable moving forwards.[9] |
| FFA→BMI | Obesity and Its Metabolic Complications: The Role of Adipokines and the Relationship between Obesity, Inflammation, Insulin Resistance, Dyslipidemia and Nonalcoholic Fatty Liver Disease.[10] |
| TG→BMI | Obesity and Its Metabolic Complications: The Role of Adipokines and the Relationship between Obesity, Inflammation, Insulin Resistance, Dyslipidemia and Nonalcoholic Fatty Liver Disease.[10] |
| Albumin→ACR | Microalbuminuria.[11] |
| Ex:HbA1c→ACR | Glycated albumin and glycated hemoglobin- A comparison.[12] |

Table 3: Digital twin map metabolic pathway reference sources

| **Pathway** | **Reference Source** |
|---|---|
| Ex:ROM→ACR | Diabetes-Induced Reactive Oxygen Species: Mechanism ofTheir Generation and Role in Renal Injury.[13] |
| Liver→Creatinine | Creatine & Creatinine Metabolism.[14] |
| Creatinine→eGFR | Prediction of creatinine clearance from serum creatinine.[15] |
| ACR→eGFR | Combining GFR and Albuminuria to Classify CKD Improves Prediction of ESRD.[16] |
| Liver+Protein→Albumin | Acute-phase proteins and other systemic responses to inflammation.[17] |
| Ex:Respiration→SBP | Effects of respiration on blood pressure and heart rate variability in humans.[18] |
| SBP→ACR | Relationship of Microalbuminuria with different Clinical and Biochemical Parameters in Newly Detected Diabetes Mellitus Cases.[19] |
| Glucose→HbA1c | Significance of HbA1c Test in Diagnosis and Prognosis of Diabetic Patients.[20] |
| Hb+ROM→HbA1c | Determination of glycated hemoglobin with special emphasis on biosensing method.[21] |
| Liver→ALT | The past and present of serum aminotransferases and the future of liver injury biomarkers.[22] |
| Liver→AST | The past and present of serum aminotransferases and the future of liver injury biomarkers.[22] |
| Ex:Nutrient→Fat-lipids | Nutrient Utilization in Humans: Metabolism Pathways.[23] |
| Ex:Nutrient→Protein | Nutrient Utilization in Humans: Metabolism Pathways.[23] |
| Ex:Nutrient→Glucose | Nutrient Utilization in Humans: Metabolism Pathways.[23] |

Table 4: GMF differences in subgroup analysis of future non-CKD vs future CKD and baseline CKD vs future CKD

| Individual GMF/Fluxes | Future non-CKD vs Future CKD | Base CKD vs Future CKD |
|---|---|---|
| Erythrocytes→Haematocrit | ↑ | ↓ |
| Ex.Respiration→Erythrocytes | ↑ | ↓ |
| Ex.Nutrient→Glucose | ↑ | ↓ |
| Albumin→ACR | ↑ | ↓ |
| Liver.Protein→Albumin | ↑ | ↓ |
| Ex.HbA1c→ACR | ↑ | ↓ |
| Liver→Creatinine | ↑ | ↓ |
| Haematocrit↔Hb | ↑ | ↓ |
| Hb.ROM→HbA1c | ↑ | ↓ |
| Ex.Nutrient→Protein | ↑ | ↓ |
| Ex.Respiration→SBP | ↑ | NA |
| Liver.Fat.lipids→HDL | NA | ↑ |
| Fat.lipids↔FFA | NA | ↓ |
| FFA→BMI | NA | ↓ |

Table 5: Comparison between the HVD model against other published predictive models for chronic kidney disease (CKD) in T2DM populations

| **Predictive models in T2DM populations** | **Outcomes (prediction horizon)** | **Model input parameter list** | **AUC (number of patients in the validation cohort)** | **Sensitivity (Sn) and Specificity (Sp)** |
|---|---|---|---|---|
| HealthVector Diabetes - Mesh Bio, Singapore | predicting the incidence of moderately to severely reduced kidney failure based on eGFR (3 years) | Age and gender plus 14 basic parameters in the GMF model (Glucose, HbA1c, Cholesterol, HDL, LDL, TG, SBP, BMI, Sr Creatinine, Sr Albumin, Haemoglobin, Haematocrit, ALT, AST) | AUC = 0.86 (n = 1,814 in testing and validation cohort) | Sn=0.80, Sp=0.74 |
| KidneyIntelx - Renalytix,[24] USA | predicting the outcome of progressive decline in kidney function using eGFR (up to 5 years) | 3 plasma biomarkers & 3 basic parameters (TNF receptors 1 and 2 (TNFR1 and TNFR2), plasma kidney injury molecule-1 (KIM-1), eGFR, uACR, and systolic BP) | AUC = 0.77 (n = 460 in validation cohort) | Sn=0.77, Sp=0.58 |
| Shanghai Diabetes Institute,[25] China | predicting the incidence of DKD (up to 4 years) | 4 basic parameters (Sex, BMI, Systolid Blood Pressure and Diabetes Duration) | AUC = 0.72 (n = 3,515 in validation cohort) | Sn=0.76, Sp=0.57 |
| Asia University,[26] Taiwan | predicting the risk of CKD in 3 time points (1, 3, & 5 years) | 11 basic parameters age, diabetes duration, insulin use, eGFR, ACR, HDL, TG, diabetes retinopathy, variation of HbA1c, variation of FPG, hypertension drug use) | AUC (average) = 0.76 (n = 1,534 in validation cohort) | Sn=0.75, Sp=0.63 |
| National Clinical Research Center for Kidney Diseases,[27] China | predicting risk of DKD (3 years) | 8 parameters (age, homocysteine, HbA1c, BMI, serum albumin, eGFR, bicarbonate, LDL) | AUC = 0.82 ( n = 164 in validation cohort) | Sn=0.78, Sp=0.77 |
| Center for Clinical Translational Innovations and Biomedical Big Data Center, Sichuan University,[28] China | predicting CKD based on eGFR and ACR (5 years) | Retinal fundus image and 6 basic parameters (age, gender, height, weight, bmi, blood pressure) | AUC = 0.72 ( n = 3,376) | Not reported |
| QKidney validation in T2DM - University Malaya,[29] Malaysia | predicting the incidence of moderate-severe DKD (5 years) | 15 basic parameters (Age, BMI, Systolic blood pressure, Smoking status, Ethnicity, Diagnosed T1DM, Diagnosed T2DM, Diagnosed RA, Diagnosed hypertension and under treatment,Diagnosed CVD, Diagnosed congestive cardiac failure, Diagnosed PVD, Diagnosed SLE, Diagnosed Renal Calculi, Positive family history of renal diseases. | AUC = 0.75 (n = 377 total patients) | Estimated Sn=0.78, Sp=0.60 |

### Figures

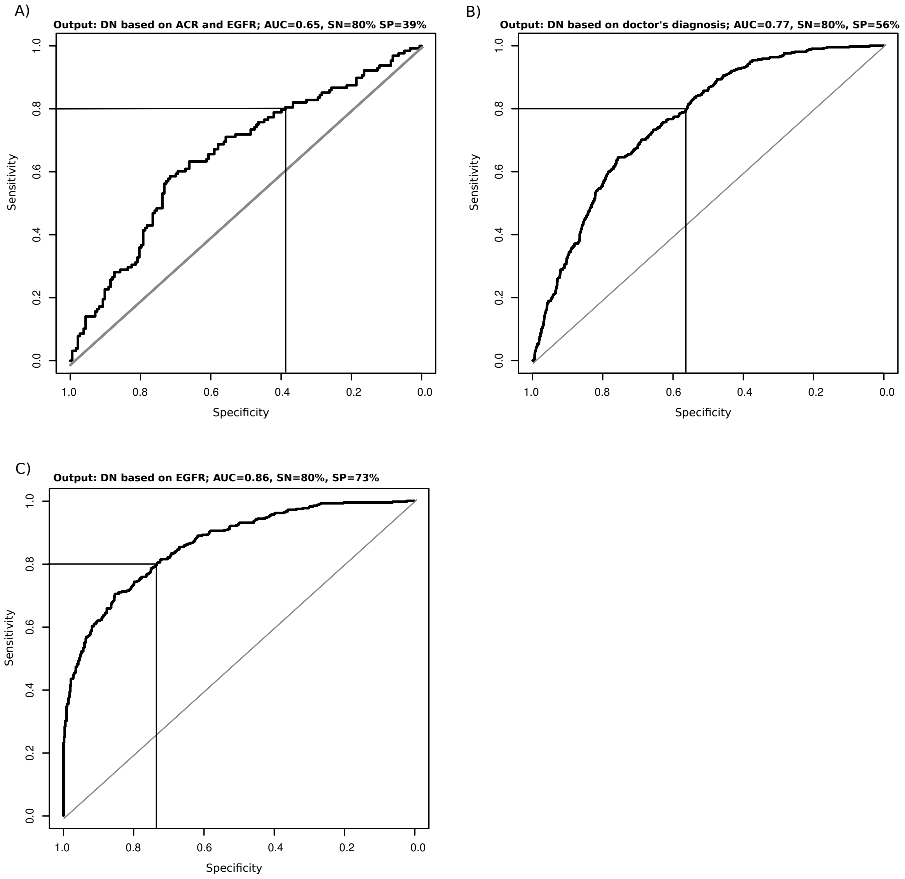


Figure 1: **Selection of eGFR as an improved endpoint for the SDR dataset.** Unlike the EVAS and the CDMD datasets, which consisted of hospital patients (the Tan Tock Seng Hosiptal), 67% of the SDR patients were from SingHealth polyclinics. In polyclinics data collection is primarily driven by the national clinical practice guidelines (CPG[30]). The Diabetes CPG recommend annual assessment of the ACR, as well as eGFR (based on serum creatinine). The CPG define the CKD status based on the eGFR values. The present study is in consistence with these quidelines. To identify the diagnostic criteria for SDR that would be most consistent with those of EVAS and CDMD, we analyzed 3 alternatives outputs for our 3-year CKD prediction model: **A**) using microalbuminuria (ACR > 3.3 mg/mMol), **B**) using the codified diagnosis record (kidney disease stage 3 or above) and **C**) eGFR < 60 mL/min/1.73. Our results indicated that using eGFR alone (**C**) provided the highest performing predictive model (AUC=0.86), followed by the diagnosis records (AUC=0.77). The observed gap between the two may be explained by the explicit CPG recommendation to estimate renal function only when eGFR is below 60 mL/min/1.73, while leaving earlier stages ambiguous.

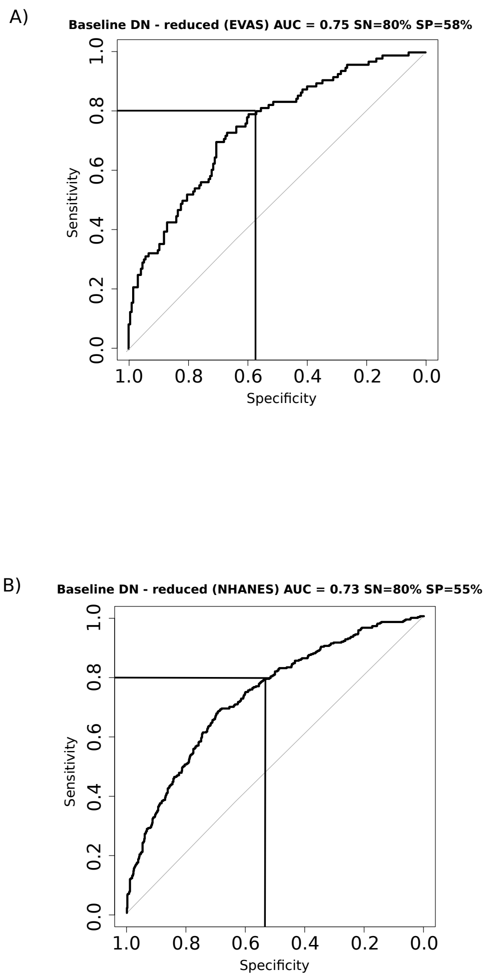


Figure 2: **Performance of identification model with a reduced set of parameters** The performance of the identification of baseline CKD model with reduced parameters is shown. The AUC achieved with the EVAS dataset was 0.75 and with the NHANES dataset was 0.73. The GMF model used here had one less input (9) parameter, serum creatinine being the excluded input.

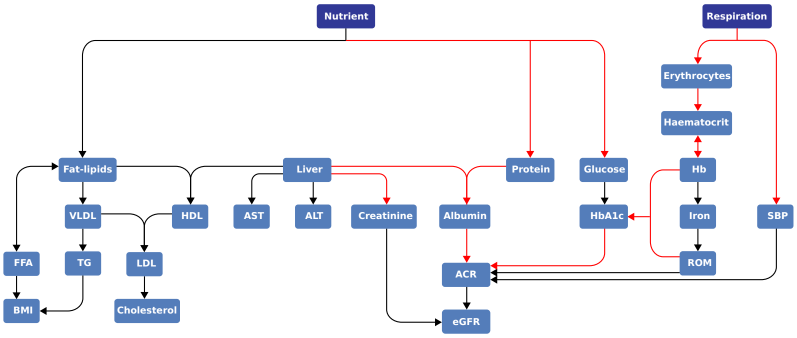


Figure 3: **GMF subgroup analysis in baseline non-CKD vs baseline CKD.** The GMF profile of patients with baseline CKD shows a poorer health profile than the patients without baseline CKD.

a)

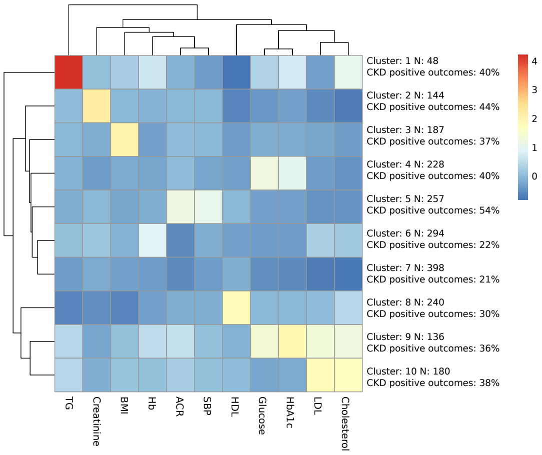


b)

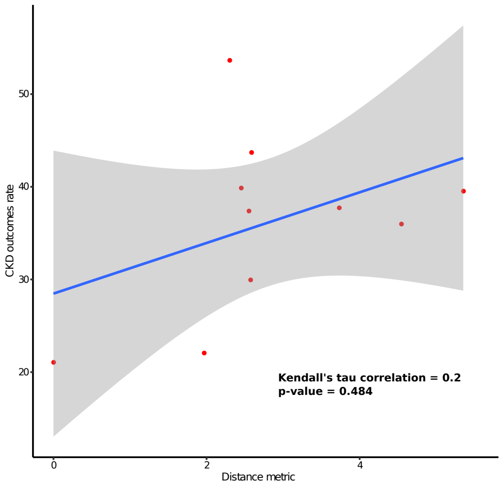


Figure 4: **Patient clustering with input parameters** The figure illustrates the clustering pattern of patients in the CDMD dataset using clinical and physiological parameters (a) and the relationship between the cluster distances and CKD outcomes rate (b). N represents the number of patients within the specific cluster, and CKD-positive outcomes are calculated as the ratio of patients developing future CKD (within 3 years) to the total number of patients in that cluster. There is no distinct pattern observed in the cluster with the highest CKD outcomes rate. There is also no significant correlation $\tau$=0.2, p=0.484) between cluster distance and CKD outcomes rate.

### References


[1] F. B. Jensen, Red blood cell pH, the bohr effect, and other oxygenation-linked phenomena in blood o2 and CO2 transport, Acta Physiologica Scandinavica 182 (3) (2004) 215–227. doi:10.1111/j.1365-201x.2004.01361.x.

[2] V. Hower, P. Mendes, F. M. Torti, R. Laubenbacher, S. Akman, V. Shulaev, S. V. Torti, A general map of iron metabolism and tissue-specific subnetworks, Molecular BioSystems 5 (5) (2009) 422. doi:10.1039/b816714c.

[3] E. Pretorius, D. B. Kell, Diagnostic morphology: biophysical indicators for iron-driven inflammatory diseases, Integrative Biology 6 (5) (2014) 486–510. doi:10.1039/c4ib00025k.

[4] G. O. Latunde-Dada, Ferroptosis: Role of lipid peroxidation, iron and ferritinophagy, Biochimica et Biophysica Acta (BBA) - General Subjects 1861 (8) (2017) 1893–1900. doi:10.1016/j.bbagen.2017.05.019.

[5] P. O. Kwiterovich, The metabolic pathways of high-density lipoprotein, low-density lipoprotein, and triglycerides: a current review, The American Journal of Cardiology 86 (12) (2000) 5–10. doi:10.1016/s0002-9149(00)01461-2.

[6] A. Jomard, E. Osto, High density lipoproteins: Metabolism, function, and therapeutic potential, Frontiers in Cardiovascular Medicine 7. doi:10.3389/fcvm.2020.00039.

[7] P. Durrington, Dyslipidaemia, The Lancet 362 (9385) (2003) 717–731. doi:10.1016/s0140-6736(03)14234-1.

[8] I. Eleftheriadou, P. Grigoropoulou, N. Katsilambros, N. Tentolouris, The effects of medications used for the management of diabetes and obesity on postprandial lipid metabolism, Current Diabetes Reviews 4 (4) (2008) 340–356. doi:10.2174/157339908786241133.

[9] S. F. Previs, D. G. McLaren, S.-P. Wang, S. J. Stout, H. Zhou, K. Herath, V. Shah, P. L. Miller, L. Wilsie, J. Castro-Perez, D. G. Johns, M. A. Cleary, T. P. Roddy, New methodologies for studying lipid synthesis and turnover: Looking backwards to enable moving forwards, Biochimica et Biophysica Acta 1842 (3) (2014) 402–413. doi:10.1016/j.bbadis.2013.05.019.

[10] U. Jung, M. Choi, Obesity and its metabolic complications: the role of adipokines and the relationship between obesity, inflammation, insulin resistance, dyslipidemia and nonalcoholic fatty liver disease, International Journal of Molecular Sciences 15 (2014) 6184–6223. doi:10.3390/ijms15046184.

[11] N. Khosla, P. A. Sarafidis, G. L. Bakris, Microalbuminuria, Clinics in Laboratory Medicine 26 (3) (2006) 635–653. doi:10.1016/j.cll.2006.06.005.

[12] B. Mukherjee, S. Patra, A. K. Das, Glycated albumin and glycated hemoglobin- a comparison, International Journal of Biomedical Research 4 (8) (2013) 381. doi:10.7439/ijbr.v4i8.301.

[13] S. Fakhruddin, W. Alanazi, K. E. Jackson, Diabetes-induced reactive oxygen species: Mechanism of their generation and role in renal injury, Journal of Diabetes Research 2017 (2017) 1–30. doi:10.1155/2017/8379327.

[14] M. Wyss, R. Kaddurah-Daouk, Creatine and creatinine metabolism, Physiological Reviews 80 (3) (2000) 1107–1213. doi:10.1152/physrev.2000.80.3.1107.

[15] D. W. Cockcroft, H. Gault, Prediction of creatinine clearance from serum creatinine, Nephron 16 (1) (1976) 31–41. doi:10.1159/000180580.

[16] S. I. Hallan, E. Ritz, S. Lydersen, S. Romundstad, K. Kvenild, S. R. Orth, Combining GFR and albuminuria to classify CKD improves prediction of ESRD, Journal of the American Society of Nephrology 20 (5) (2009) 1069–1077. doi:10.1681/asn.2008070730.

[17] C. Gabay, I. Kushner, Acute-phase proteins and other systemic responses to inflammation, New England Journal of Medicine 340 (6) (1999) 448–454. doi:10.1056/nejm199902113400607.

[18] J.-L. Elghozi, D. Laude, A. Girard, Effects of Respiration on Blood Pressure and Heart Rate Variability in Humans, Clinical and Experimental Pharmacology and Physiology 18 (11) (1991) 735–742. doi:10.1111/j.1440-1681.1991.tb01391.x.

[19] I. Prasad, Z. Latif, T. Ahmed, F. Pathan, S. Ashrafuzzaman, F. Amin, Relationship of microalbuminuria with different clinical and biochemical parameters in newly detected diabetes mellitus cases, Ibrahim Medical College Journal 4 (2010) 21. doi:10.3329/imcj.v4i1.5931.

[20] S. I. Sherwani, H. A. Khan, A. Ekhzaimy, A. Masood, M. K. Sakharkar, Significance of HbA1c test in diagnosis and prognosis of diabetic patients, Biomarker Insights 11 (2016) BMI.S38440. doi:10.4137/bmi.s38440.

[21] C. S. Pundir, S. Chawla, Determination of glycated hemoglobin with special emphasis on biosensing methods, Analytical Biochemistry 444 (2014) 47–56. doi:10.1016/j.ab.2013.09.023.

[22] M. R. McGill, The past and present of serum aminotransferases and the future of liver injury biomarkers, Experimental and Clinical Sciencesdoi:10.17179/EXCLI2016-800.

[23] A. T. Da Poian, T. El-Bacha, M. R. Luz, Nutrient utilization in humans: Metabolism pathways, Nature Education 5 (8) (2010) 11.

[24] L. Chan, G. N. Nadkarni, F. Fleming, J. R. McCullough, P. Connolly, G. Mosoyan, F. E. Salem, M. W. Kattan, J. A. Vassalotti, B. Murphy, M. J. Donovan, S. G. Coca, S. M. Damrauer, Derivation and validation of a machine learning risk score using biomarker and electronic patient data to predict progression of diabetic kidney disease, Diabetologia 64 (7) (2021) 1504–1515. doi:10.1007/s00125-021-05444-0.

[25] M. Wu, J. Lu, L. Zhang, F. Liu, S. Chen, Y. Han, F. Zhao, K. Guo, Y. Bao, H. Chen, W. Jia, A non-laboratory-based risk score for predicting diabetic kidney disease in chinese patients with type 2 diabetes, Oncotarget 8 (60) (2017) 102550–102558. doi:10.18632/oncotarget.21684.

[26] C.-C. Lin, M. J. Niu, C.-I. Li, C.-S. Liu, C.-H. Lin, S.-Y. Yang, T.-C. Li, Development and validation of a risk prediction model for chronic kidney disease among individuals with type 2 diabetes, Scientific Reports 12 (1). doi:10.1038/s41598-022-08284-z.

[27] W. Dong, E. Y. F. Wan, D. Y. T. Fong, R. L. P. Kwok, D. V. K. Chao, K. C. B. Tan, E. M. T. Hui, W. W. S. Tsui, K. H. Chan, C. S. C. Fung, C. L. K. Lam, Prediction models and nomograms for 10-year risk of end-stage renal disease in chinese type 2 diabetes mellitus patients in primary care, Diabetes, Obesity and Metabolism 23 (4) (2021) 897–909. doi:10.1111/dom.14292.

[28] K. Zhang, X. Liu, J. Xu, J. Yuan, W. Cai, T. Chen, K. Wang, Y. Gao, S. Nie, X. Xu, X. Qin, Y. Su, W. Xu, A. Olvera, K. Xue, Z. Li, M. Zhang, X. Zeng, C. L. Zhang, O. Li, E. E. Zhang, J. Zhu, Y. Xu, D. Kermany, K. Zhou, Y. Pan, S. Li, I. F. Lai, Y. Chi, C. Wang, M. Pei, G. Zang, Q. Zhang, J. Lau, D. Lam, X. Zou, A. Wumaier, J. Wang, Y. Shen, F. F. Hou, P. Zhang, T. Xu, Y. Zhou, G. Wang, Deep-learning models for the detection and incidence prediction of chronic kidney disease and type 2 diabetes from retinal fundus images, Nature Biomedical Engineering 5 (6) (2021) 533–545. doi:10.1038/s41551-021-00745-6.

[29] Y. Sheng Qian, F.-M. Moy, Predicting the risk of chronic kidney disease among type 2 diabetes mellitus patients in a primary care setting: An evaluation of the qkidney model, Malaysian Journal of Medicine and Health Sciences 15 (2019) 67–73.

[30] Clinical practice guidelines. diabetes mellitus., techreport, Ministry of Health, Singapore, College of Medicine Building, 16 College Road, Singapore 169854 (2014). URL http://www.moh.gov.sg/cpg
